# Magnitude and Predictors of Anemia Among Adult Solid Cancer Patients in Hawassa, Ethiopia: A Cross-Sectional Study

**DOI:** 10.64898/2026.01.21.26344582

**Authors:** Eden Alemu, Abebe Melis, Alemwosen T/haimanot, Bizunesh Dires, Anteneh Fikrie, Hagos Tesfay, Abdela Amano

**Author notes:** Email of authors Eden Alemu Abebe Melis Alemwosen T/haimanot Bizunesh Dires Anteneh Fikrie Hagos Tesfay Abdela Amano.

## Abstract

**Introduction:** Anemia is a common and debilitating complication in cancer patients, affecting prognosis, quality of life, and treatment response. The burden of anemia may be higher in resource-limited settings due to nutritional deficiencies and limited healthcare access. However, data on anemia among newly diagnosed cancer patients in Ethiopia remain scarce. This study was aimed to determine the magnitude and predictors of anemia in newly diagnosed adult solid cancer patients in Hawassa, Ethiopia.

**Methods:** A cross-sectional study was conducted among randomly selected 405 newly diagnosed adult solid cancer patients at selected public health facilities in Hawassa City, Ethiopia, from June 2021 to October 2021. Data were collected using structured face-to-face interviews and from medical records. Data were entered into EpiData version 3.1 and exported to SPSS version 26 for analysis. Descriptive statistics were computed for all variables. Binary logistic regression was performed to determine the independent effects of potential explanatory variables on the presence of anemia. Adjusted Odds ratios with 95% confidence intervals were calculated. Statistical significance was set at p-value < 0.05.

**Result:** The prevalence of anemia was 26.4% (95% CI: 22.1–30.7%), with normocytic anemia (56.1%) being the most common morphological type. Multivariable analysis identified a history of bleeding (AOR = 3.46, 95% CI: 1.94–6.16), advanced cancer stage (AOR = 2.01, 95% CI: 1.20–3.34), and underweight BMI (AOR = 2.14, 95% CI: 1.28–3.55) were significantly associated with anemia. Conversely, higher educational attainment (diploma and above) was protective (AOR = 0.35, 95% CI: 0.13–0.93).

**Conclusion:** Anemia affects over one-quarter of newly diagnosed solid cancer patients in Hawassa, with advanced-stage disease, bleeding history, and decreasing body mass index as significant risk factors. These findings highlight the need for routine anemia screening at diagnosis, nutritional interventions, and early bleeding management in oncology settings.

## Introduction

Cancer is a major public health concern worldwide, characterized by the uncontrolled proliferation and spread of abnormal cells that can invade surrounding tissues, metastasize to distant organs, and result in death(1,2). Globally, cancer caused 10 million deaths in 2020, with 19.3 million new cases(3). In Ethiopia and other low- and middle-income countries (LMICs), cancer burden is rising due to aging, increased risk factors, and poor access to early detection and treatment(4).

Anemia is a common and debilitating complication in cancer patients, significantly impacting prognosis, quality of life, and treatment response(5). It is characterized by reduced hemoglobin (Hb) concentration, red blood cell (RBC) count, or packed cell volume, leading to impaired oxygen delivery to tissues(6). The National Comprehensive Cancer Network (NCCN) classifies anemia as mild (Hb 10–lower limit of normal g/dL), moderate (Hb 8–<10 g/dL), severe (Hb 6.5– <8 g/dL), and life-threatening (Hb <6.5 g/dL)(7). Cancer-related anemia (CRA) is multifactorial, influenced by tumor-induced inflammation, chemotherapy-induced myelosuppression, blood loss, nutritional deficiencies, and chronic disease states(8,9). Based on RBC morphology, CRA can be microcytic (commonly due to iron deficiency), normocytic (linked to chronic disease), or macrocytic (resulting from vitamin B12 and folic acid deficiencies)(9,10).

The prevalence of anemia in cancer patients varies depending on tumor type, disease stage, treatment modality, and geographical region. The European Cancer Anemia Survey (ECAS) reported a prevalence of 39.3%(11), while rates in China and Saudi Arabia were 63.2% and 44.1%, respectively(12,13). In sub-Saharan Africa, anemia remains a major comorbidity among cancer patients, exacerbated by factors such as delayed diagnosis, limited access to treatment, and high prevalence of infectious diseases. In Tanzania, 38.6% of cervical cancer patients presented with severe anemia, reflecting the substantial burden of the condition in resource-limited settings(14). Similarly, Ethiopian studies have reported anemia prevalence ranging from 23% to 54.8% across different solid tumors, with the highest rates observed in patients with advanced-stage disease(15,16). Among specific tumor types, breast cancer patients had a prevalence of 21.7%, while cervical cancer patients exhibited a significantly higher rate of 50.95%(10).

Despite the growing recognition of CRA as a critical issue in oncology, data on its prevalence and associated factors among newly diagnosed solid cancer patients in Ethiopia remain scarce. Most studies conducted in Africa have focused on specific cancer types or included patients already undergoing treatment, making it difficult to assess anemia at baseline. Understanding the burden and determinants of anemia in newly diagnosed cancer patients is essential for developing targeted interventions aimed at improving patient outcomes.

This study assessed the magnitude, morphological types, and associated factors of anemia among newly diagnosed adult solid cancer patients attending oncology services in Hawassa city health facilities, Ethiopia. The identified key predictors such as cancer stage, nutritional status, and bleeding history, can be used to practice evidence-based strategies for early detection and management of anemia in cancer care settings.

## Methods

### Study area and design

The study was conducted at government and private health facilities in Hawassa city, which the capital city of newly structured Sidama regional state. The city is located 275 km to southeast of Addis Ababa. It has divided in to 8-sub city administration and with estimated population of 350,000; it covers 50 km^2^ and is located 1,700m above sea level. Hawassa University Comprehensive Specialized Hospital (HUCSH) is the first referral hospital established in the region. It serves as the teaching hospital at Hawassa University College of Medicine and Health Sciences (HU-CMHS), it is serving more than 18 million people of the Sidama region, Southern Peoples Regional State, and the surrounding Oromia region zones.

HU-CMHS offers undergraduate, postgraduate, PhD and specialty programs in various departments, including Health Officer, Nursing, Pharmacy, Medicine, Optometry, General Public Health, Epidemiology, Reproductive Health, PhD in Public Health, Internal Medicine, Surgery, Gynecology and Obstetrics, Pediatrics, Pathology, and Ophthalmology. Cytology and histopathology services are available at the pathology departments of HUCSH, Yanet Specialized Medical Center, and Alatyon General Hospital. Cancer diagnosis is conducted through both cytological and histopathological examinations. These three healthcare facilities provide diagnostic services and cancer therapy.

### Study Design and Period

A health facility based cross sectional study was conducted on solid cancer adult patients from 1^st^ June to 30^th^ October, 2021.

### Population, Eligibility, Sample size determination and sampling technique and procedure

The study population comprised all newly diagnosed adult patients with solid cancers receiving outpatient or inpatient care at the following health facilities in Hawassa City: HUCSH, Yanet Specialized Medical Center, and Alatyon General Hospital. Inclusion criteria included individuals aged 18 years or older, newly diagnosed with a solid tumor confirmed by histopathology or cytology, and willing to provide written informed consent. Exclusion criteria encompassed patients with hematological malignancies (e.g., leukemia), terminal illnesses, severe cognitive impairment, or inability to provide written informed consent, as well as those who had received blood transfusions or iron supplementation within two weeks prior to enrollment.

The sample size was determined using Epi Info software, assuming a 95% confidence level, 80% power, and a prevalence of anemia among cancer patients of 23%(15). A 10% non-response rate was also considered. This resulted in a calculated sample size of 405 participants. Hawassa City has five governmental hospitals, five private hospitals, and over 60 clinics, including one Specialized Medical Center. We selected HUCSH (governmental), Alatyon General Hospital (private), and Yanet Specialized Medical Center (private) based on their availability of cytology, histopathology, and oncology services, ensuring adequate representation of newly diagnosed solid cancer patients. Eligible patients were identified from the oncology departments of these facilities using their cytology/histopathology results. The sample size was proportionally allocated across the three facilities. All eligible patients presenting to the oncology departments during the study period were selected systematically.

### Study variables, operational definitions

The dependent variable was magnitude of anemia. Whereas the independent variables were Socio-demographic factors: Age, sex, marital status, occupational status, educational status, residency, BMI, type of cancer, stage of cancer, and bleeding history.

#### Solid cancer

A type of cancer that forms a discrete mass and occurs in solid organs such as the kidney, breast, cervix, colon, etc., excluding leukemia(1).

#### Cancer related anemia

Defined as hemoglobin levels less than 11 g/dl(17).

#### Underweight

Defined as a body mass index (BMI) of less than 18.5 kg/m^2^.

### Data collection tools and procedures and data quality management

Data were collected by three trained data collectors using a standardized data collection instrument. A data abstraction checklist was developed based on a review of relevant literature and patient medical records. This checklist was used to extract clinical findings, complete blood count results, and other relevant information from patient medical charts. The data collection tool consisted of sociodemographic characteristics, clinical findings, and complete blood and anthropometric measurements. Height and weight were measured using calibrated measuring tapes and weighing scales, respectively. Body Mass Index (BMI) was calculated from these measurements. Data quality was ensured through regular supervision and data cleaning activities by the research team. Inconsistent or missing values were verified with patient records before analysis.

### Data processing and analysis

Data were coded and entered into EpiData version 3.1 and then exported to SPSS version 26 for analysis. Data were cleaned, and checked for missing values and outliers. Descriptive statistics (frequencies, means, standard deviations, and percentages) were calculated for all variables. Tables and graphs were used to present the descriptive findings. Chi-square tests were used to examine the association between anemia and categorical variables. Bivariable and multivariable binary logistic regression was performed to assess the independent effect of each explanatory variable on anemia, while controlling for other confounding factors. Variables with a p-value < 0.25 in the bivariable analysis were included in the multivariable model. The Hosmer-Lemeshow test was used to assess the goodness-of-fit of the final logistic regression model. Odds ratios (ORs) with 95% confidence intervals (CIs) were calculated to quantify the strength of the association between independent variables and anemia.

### Ethical consideration

Ethical clearance and approval of the study was obtained from institutional review board (IRB) of Hawassa University College of Medicine and Health Science with reference number IRB/182/13. Verbal and written informed consent were obtained from all respondents before each interview. The consent was translated to local language and read for participants. The confidentiality of the study participants was entirely kept secret. The respondents were informed about the objective and purpose of the study and they were also told that they could quit the study at any time they wanted. Personal privacy and dignity were respected. Clinical information was abstracted from patient medical records during the study period and access to these records was limited to the research team, and no identifiable personal information was retained in the dataset used for analysis.

## Result

### Socio-demographic characteristics

Out of the 405 respondents, 59.8% were female. The majority (39.3%) were in the 35–49-year age group, with an age range of 18 to 85 years and a mean age of 46.29 years (SD ±13.78). More than half (52.1%) resided in urban areas, and 82.2% were married. Regarding education, 34.1% had no formal education, 28.4% had completed primary education, 19.8% had secondary education, and 17.8% held a diploma and above. In terms of occupation, 45.9% were unemployed, followed by 31.9% who were employed and 22.2% who were farmers. (Table1).

**Table 1:**
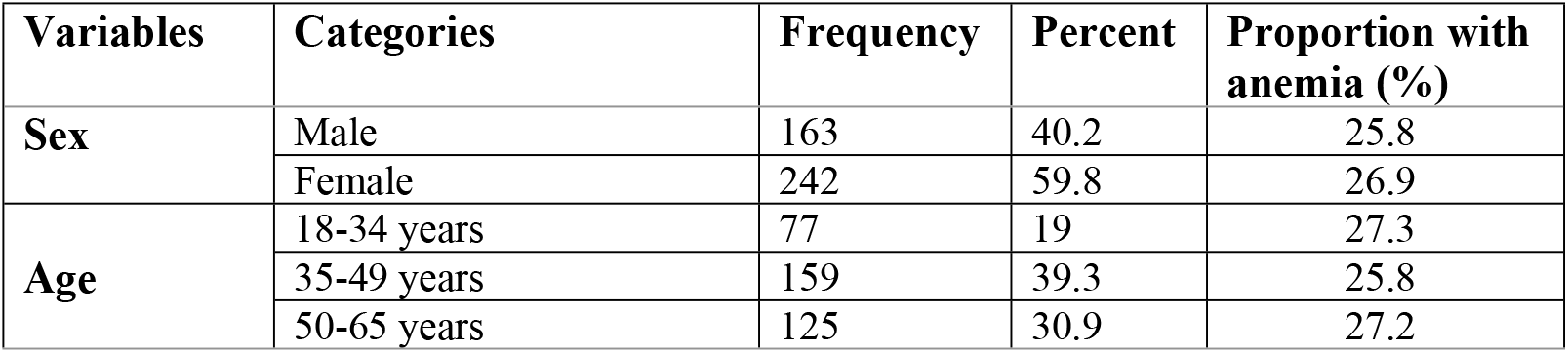
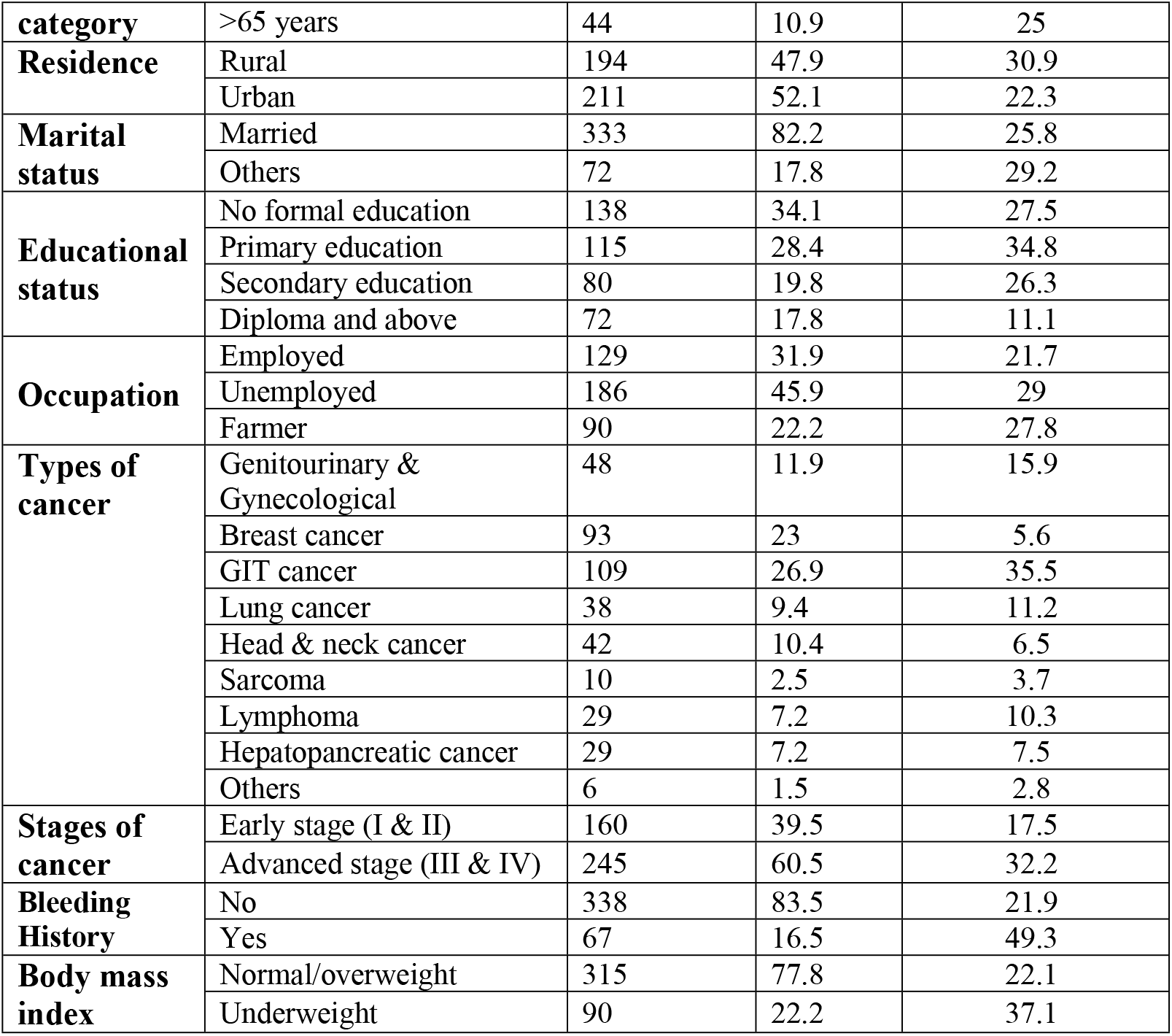
Socio-demographic and clinical characteristics associated with anemia among adult solid cancer patients in Hawassa, Ethiopia (N=405), 2022.

### Magnitude of anemia

The mean hemoglobin concentration among participants was 12.15 g/dl (SD ±2.27), with an overall anemia prevalence of 26.4% (95% CI: 22.3%, 30.9%). A notably higher prevalence of anemia (35.5%) was observed in individuals with gastrointestinal cancers. Among cancer types, 26.9% had gastrointestinal cancer and 23% had breast cancer. More than half of the patients (60.5%) were diagnosed at an advanced stage (III & IV), among whom the prevalence of anemia was 32.2%.

A history of bleeding was reported by 16.5% of participants, with vaginal bleeding accounting for 34.3% of these cases. Among those with bleeding history, 49.3% were anemic. Despite this, 57.9% of participants had no treatment plan for anemia. Among those with a treatment plan, 62.2% received blood transfusions and 37.8% were prescribed iron supplements.

The participants had a mean BMI of 20.6kg/m2 (SD ±3.71), with values ranging from 13.5 to 35.3. About 22.2% of participants were classified as underweight, and 37.1% of them were anemic. (Table1).

### Morphologic types of anemia

The participants in this study had a MCV of 84.2 ± 7.9 fl. Based on MCV, 56.1% of the anemic participants had normocytic anemia, 43% had microcytic anemia,, and only 0.9% had macrocytic anemia (Figure 1).

**FIGURE 1:**
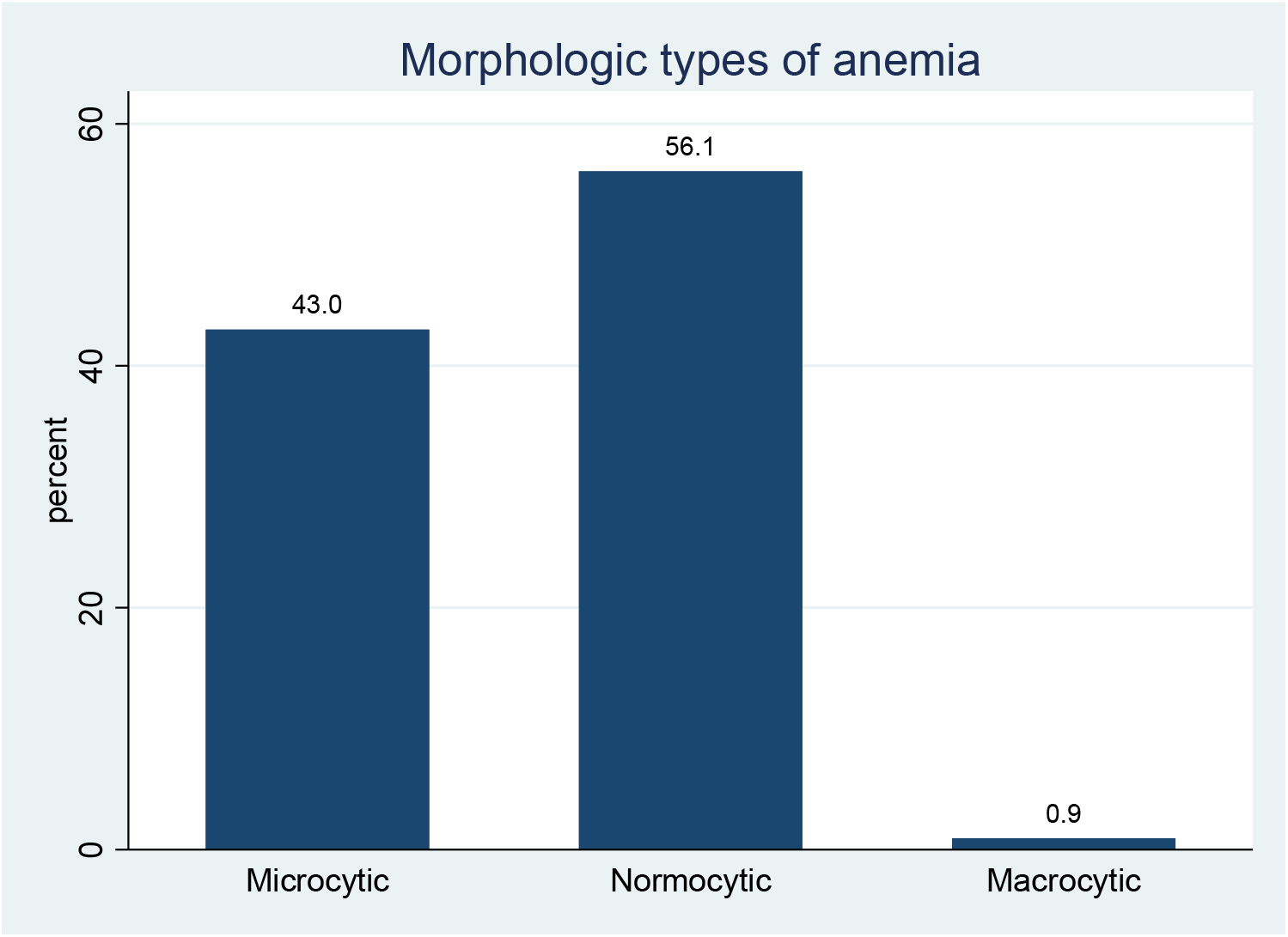
Morphologic types of anemia among newly diagnosed solid cancer adult patients in Hawassa city health facilities, Hawassa, Ethiopia (N=107), 2022 *Microcytic: MCV<80fl, Normocytic: 80fl* ≤ *MCV*≥*100fl, and Macrocytic: MCV >100fl*.

### Predictors of anemia

In a bivariate analysis; place of residence, educational status, stage of cancer, bleeding history, and body mass index of the participants were associated with anemia. In multivariable analysis educational status, stage of cancer, bleeding history, and body mass index remained significantly associated with the occurrence of anemia. (Table 2).

**Table 2:** Bivariable and Multivariable Predictors of Anemia Among Adult Solid Cancer Patients in Hawassa, Ethiopia (N=405), 2022.

| Variables | Categories | Crude OR [95% CI] | P-value | Adjusted OR [95% CI] | P-value |
| --- | --- | --- | --- | --- | --- |
| Residence | Urban | 0.64 [0.41–0.99] | 0.049* | 0.71 [0.4–1.26] | 0.249 |
|  | Rural | 1 | — | 1 | — |
| Educational status | Primary Education | 1.40 [0.82–2.40] | 0.215 | 1.51 [0.85–2.69] | 0.16 |
|  | Secondary Education | 0.94 [0.50–1.75] | 0.837 | 1.08 [0.53–2.2] | 0.82 |
|  | Diploma and above | 0.33 [0.14–0.75] | 0.008* | 0.35 [0.13–0.93] | 0.035** |
|  | No formal education | 1 | — | 1 | — |
| Occupation | Unemployed | 1.48 [0.87–2.49] | 0.146 | 0.71 [0.35–1.4] | 0.33 |
|  | Farmer | 1.39 [0.74–2.59] | 0.303 | 0.52 [0.22–1.21] | 0.13 |
|  | Employed (Ref) | 1 | — | 1 | — |
| Bleeding history | Yes | 3.46 [2.01–5.97] | 0.001* | 3.46 [1.94–6.16] | 0.001** |
|  | No | 1 | — | 1 | — |
| BMI | Underweight | 2.07 [1.30–3.31] | 0.002* | 2.14 [1.28–3.55] | 0.003** |
|  | Normal/overweight | 1 | — | 1 | — |
| Cancer stage | Advanced stage | 2.24 [1.38–3.65] | 0.001* | 2.00 [1.2–3.34] | 0.008** |
|  | Early stage | 1 | — | 1 | — |

NB. Statistically significant associations are marked with \* (Crude OR) and \*\* (Adjusted OR). The model demonstrated good fit (Hosmer–Lemeshow $\chi^2 = 5.99$ , $p = 0.65$ ).

Participants with a history of bleeding had 3.46 times increased odds of anemia (AOR = 3.46, 95% CI: 1.94–6.16). Similarly, patients with advanced-stage cancer had double the odds of being anemic compared to those with early-stage disease (AOR = 2.00, 95% CI: 1.2–3.34). BMI was another significant predictor: underweight patients were twice as likely to be anemic compared to those with normal or overweight BMI (AOR = 2.14, 95% CI: 1.28–3.55). Educational attainment was protective: participants with a diploma or higher had significantly lower odds of anemia compared to those with no formal education (AOR = 0.35, 95% CI: 0.13–0.93).

## Discussion

This study found an overall anemia prevalence of 26.4% among newly diagnosed adult solid cancer patients in Hawassa. This figure is comparable to the prevalence reported in Addis Ababa 23% (15), but slightly higher than rates observed in China, 18.9%(12). Conversely, our report is lower than the findings made by other researchers that revealed 44.1% in Saudi Arabia (13), and 64% India (19). Such differences may reflect variations in the definition of anemia, study population, cancer types, and nutritional status across settings.

A higher prevalence of anemia was observed in patients with gastrointestinal cancers (35.5%), consistent with the role of chronic, often occult bleeding in digestive tract malignancies. Our findings are lower compared to other studies which reported 77.5% for gynecological and 67.9% GIT cancers(9).

Regarding the anemia treatment plan, our findings revealed that anemia was planned to be treated in 42.1% of patients with anemia. This result was similar to the reports conducted in Australian Cancer Anemia Survey (20)and European Cancer Anemia Survey (11) which was done in 24 European countries showed, 41%% and 38.9% of patients with anemia were treated for their anemia before receiving any anti-cancer therapy, respectively, however our finding was higher than Addis Ababa study (15) in which 32% of patients with anemia got anemia correction treatment prior to commencing anti-cancer treatment. The most common treatment for anemia correction was blood transfusion (62.2%) in the current study, which is higher than that of ACAS (20) which was 36%. Majority of patients in our survey had mild to moderate type of anemia which was 82.3%, which agrees with that of Indian study which reported 92%(21).

The current study showed that 56.1% of the anemic patients had normocytic anemia, 43% had microcytic anemia and only 0.9% had macrocytic anemia. A similar study conducted in the United States reported normocytic anemia in 68% of cases, microcytic anemia in 31%, and macrocytic anemia in 2% (18). In contrast, an Indian study found iron deficiency to be the most common etiology of anemia in patients with solid tumors (54.33%), followed by anemia of chronic disease (41.73%), while megaloblastic anemia was observed in 3.94% of cases (19). These differences may be attributed to variations in study population size and differences in nutritional status across regions.

Multivariable analysis identified bleeding history, advanced cancer stage, educational status and low BMI as independent predictors of anemia. Patients with bleeding history had more than threefold increased odds of anemia. The possible justification for this could be patients with bleeding history have a higher risk for anemia occurrence and this was also confirmed in our finding that revealed about half of the patients (49.3%) who had bleeding history were anemic, this finding agreed with the study done in China (12).

Advanced-stage cancers doubled anemia risk, likely due to delayed presentation and increased tumor burden, although studies from China(12) and Italy(23) did not find stage to be significant. High educational attainment was protective, with diploma-level participants significantly less likely to be anemic, suggesting that health literacy and care-seeking behaviors may mitigate anemia risk. This finding is consistent with evidence from Ethiopia, where schooling has been shown to reduce anemia prevalence(24).

Cancer patients who had lower body mass index were two times more likely to be anemic than patients who had higher body mass index. This could be because of cancers may lead to decreased food intake by varieties of mechanisms, such as anorexia, nausea, and other factors increased basal metabolic rate, obstruction in the digestive tract, maldigestion, diarrhea or constipation, and with so many reasons, which finally lead to poor nutrition absorption and malnutrition that may result low BMI eventually leads anemia. Our report is consistent with study done in Italy (9) that showed a significant positive relationship was found between Hb and BMI.

The strength of this study was, since no similar study was done in this study setting, the study finding may serve as a base line data for other researchers and physicians. Despite these findings, this study was limited by its cross-sectional design, which prevents causal inferences. Future longitudinal studies should assess anemia progression over time

## Conclusions

Anemia affects 26.4% of newly diagnosed solid cancer patients in Hawassa, with bleeding history, advanced stage, and decreasing BMI as significant risk factors. These findings highlight the need for comprehensive anemia screening, early intervention, and nutritional support in Ethiopian oncology care.

## Recommendations

➢ Routine anemia screening at cancer diagnosis.
➢ Early bleeding management for high-risk cancers (e.g., gastrointestinal, gynecologic).
➢ Nutrition-based interventions targeting iron, B12, and folate deficiencies.
➢ Research: - Further studies on absolute and functional iron deficiency in solid cancer patients, and parasite exclusion through stool examination -Future studies should assess the impact of anemia treatment on cancer outcomes, including survival and treatment response, to guide clinical decision-making

## Data Availability

All data underlying the findings described in this manuscript are fully available without restriction. The complete dataset supporting the conclusions of this study has been deposited and is accessible through the corresponding author, Dr. Abebe Melis Nisro, at Hawassa University Comprehensive Specialized Hospital. Data are available from the time of publication and can be accessed without limitation.

## Data Availability

The data used to support the findings of this study are available from the corresponding author upon reasonable request.

